# Pediatric Neural Changes to Physical and Emotional Pain After Intensive Interdisciplinary Pain Treatment: A Pilot Study

**DOI:** 10.1101/2023.10.03.23295921

**Authors:** Rebecca J Lepping, Cara M Hoffart, Amanda S Bruce, Jasmine M Taylor, Neil J Mardis, Seung-Lark Lim, Dustin P Wallace

## Abstract

Brain areas activated during pain can contribute to enhancing or reducing the pain experience, showing a potential connection between chronic pain and the neural response to pain in adolescents and youth. This study examined changes in brain activation associated with experiencing physical pain, and the observation of physical and emotional pain in others, by using functional magnetic resonance imaging (fMRI) before and after intensive interdisciplinary pain treatment (IIPT). Eighteen youth (age 14 to 18) with widespread chronic pain completed fMRI testing before and after IIPT to assess changes in brain activation in response to physical and emotional pain. Broadly, brain activation changes were observed in frontal, somatosensory, and limbic regions. These changes suggest improvements in descending pain modulation via thalamus and caudate, and the different pattern of brain activation after treatment suggests better discrimination between physical and emotional pain. Brain activation changes were also correlated with improvements in clinical outcomes of catastrophizing (reduced activation in right caudate, right mid-cingulate, and postcentral gyrus) and pain-related disability (increased activation in precentral gyrus, left hippocampus, right middle occipital cortex, and left superior frontal gyrus). These changes support interpretation that reduced brain protective responses to pain were associated with treatment-related improvements. This pilot study highlights the need for larger trials designed to better understand the brain mechanisms involved in pediatric widespread pain treatment.

**Perspective:** This study examined changes in brain response to pain in youth experiencing chronic pain participating in intensive interdisciplinary pain treatment (IIPT). The novelty in this pilot study is the longitudinal neuroimaging findings in conjunction with established, clinically relevant behavioral and self-report measures of pain.

## Introduction

Chronic pain lasting over three months [1] can negatively affect quality of life for youth and lead to significant disability. Between 25-37% of youth report continuous pain that is moderately severe to disabling for 5-8% [2, 3]. Forty percent exhibited fear-related pain leading to symptoms of anxiety, depression, and avoidant behavior [4, 5], implying a link between physical and emotional pain and how the two are neurologically interpreted. The fear-avoidance model was initially focused on low back pain [6] and extended to neuropathic pain and complex regional pain syndrome among others [3]. This model shows that pain can be interpreted as nonthreatening or catastrophic, where pain hinders participation in activities one enjoys [7]. Viewing pain as catastrophic can lead to a fear of pain, mobility issues, and increased fear of physical activities [3, 8].

Brain regions – including primary somatosensory cortex, primary and supplementary motor cortices, and thalamus – are responsible for processing physical pain [9]. These regions which are activated during physical pain are also activated by viewing others experiencing pain [10, 11]. Brain regions underlying fear, anxiety, and memory processing can increase the experience of pain [10]. Further, neural underpinnings of the brain’s interpretation of pain show a relationship between the physical presence of pain, pain-related fear, and how this fear can create a mental protective mechanism [7, 10]. Chronic pain can thus create a positive feedback loop whereby increasing physical and emotional pain leads to further impairment and avoidance, further increasing both pain and fear of pain (including catastrophizing).

Treatment of chronic pain integrates exercise (often supervised by physical or occupational therapists) with psychological treatments. Treatments may include exposure to discomfort by participating in certain exercises and engaging in purposeful desensitization (stimulating the skin in ways that are safe but cause discomfort). Treatments also address bodily stress systems which have become dysregulated by ongoing pain and pain-related fear. For young people highly impaired by chronic pain, intensive interdisciplinary pain treatment (IIPT) provides an effective, rehabilitation-focused approach to learning to function during pain and can lead to improvement in pain and other symptoms [12–14]. IIPT typically includes 6-8 hours per day of treatment integrating physical and occupational therapy, music and art therapy, psychological therapies, and mindfulness and relaxation-based self-regulation. Treatment is typically delivered 5 days per week, over the course of 3-6 weeks.

As IIPT includes exposure to pain as well as feared movements and other activities, it is hypothesized to decrease the brain’s protective responses. However, this has not been directly investigated in young people with widespread pain undergoing intensive treatment of this nature. It is not known to what extent IIPT would result in changes in brain response to experiencing pain and witnessing others experiencing pain, or whether this treatment could lead to changes in brain responses to emotional pain.

This longitudinal neuroimaging pediatric pain pilot study examined brain responses of adolescents with widespread pain while experiencing physical pain and observing pictures of others experiencing physical and emotional pain, prior to intensive treatment. Participants then participated in IIPT for 3-6 weeks, based on individual treatment need, and repeated the neuroimaging protocol. This study tested the hypothesis that there will be observable changes in the adolescent’s brain response and ability to decipher between experiencing physical and emotional pain after intensive treatment. As the intervention concluded, we hypothesized a difference between participants’ interpretation of physical and emotional pain, with greater reduction in brain activation to physical pain after the treatment program. This exploration of the brain’s neurological response to pain in adolescents provides the opportunity to tailor treatments for adolescents experiencing chronic pain and pain-related fear.

## Methods

### Participants

This study was conducted in a large children’s hospital in the Midwest United States. The Institutional Review Board (IRB) of Children’s Mercy Hospital gave ethical approval for this work. Adolescents ages 14-18 years with widespread chronic pain who were enrolled in an intensive interdisciplinary pain treatment (IIPT) program in the day hospital were invited to participate in this research study by clinical staff. Youth who were unable to participate in imaging due to developmental delay, implanted devices unsafe for the MR environment (i.e., metal implants or devices), or difficulty remaining still during imaging procedures were excluded from participation. Interested potential participants were then contacted by research staff who reviewed the study procedures with the participant and parent and answered any questions. Minor assent and parental permission (for those under 18 years), or consent from those 18 years and over was obtained at the participant’s first neuroimaging research visit.

## Materials

Questionnaires were administered before and after the IIPT program including the Fear of Pain Questionnaire – Child and Parent Report (FOPQ-C/FOPQ-P), the Widespread Pain Index (WPI), the Functional Disability Index (FDI), and Catastrophizing. Pain ratings were also obtained using a 100mm Visual Analog Scale (VAS) at several points throughout the study.

The Fear of Pain Questionnaire, Child Report (FOPQ-C) contains 24 self-assessment questions with two subscales: fear of pain and avoidance of activities [15]. Participants recorded any pain experienced for a few hours or days. Scores ranged on a 5-point Likert scale from 0 = ‘strongly disagree’ to 4 = ‘strongly agree’. Parents assessed the participant’s pain-related fear using the Fear of Pain Questionnaire, Parent Report (FOPQ-P). The FOPQ-P contains 23 questions with three subscales: fear of pain, avoidance of activities, and school avoidance [15]. Responses were recorded on a 5-point Likert scale from 0 = ‘strongly disagree’ to 4 = ‘strongly agree’. FOPQ reports were scored by summing the subscale scores and all responses for an overall scale score.

Participants self-assessed pain using the Widespread Pain Index (WPI), a survey containing a front/back diagram of 19 body areas to measure painful body regions [16]. Participants completed 3 categorical scales indicating symptoms over the past week for fatigue, waking up tired (unrefreshed), and trouble thinking or remembering. Level of symptom severity ranged from ‘no problem’ to ‘severe problem: pervasive, continuous, life disturbing’.

The Functional Disability Inventory (FDI) rates participants’ difficulty in functioning related to their physical health [17]. The instrument provides 15 questions to document activity limitations in the past two weeks: higher scores indicating greater disability. Total scores are calculated by combining the total sum of each rating.

Catastrophizing was assessed with the 5-item catastrophizing subscale of the Pain-Related Cognitions Questionnaire for Children (PRCQ-C) [18]. Using the item stem “when I am in pain,” the measure asks patients to reflect on what they typically think and feel when experiencing pain and then provide a frequency rating on a 5-point Likert-type scale which ranges from (1) Never to (5) Very Often. Subscale scores are calculated as an average frequency rating, with higher scores indicating greater frequency of that type of cognition.

### Procedures

Participants underwent two MRI scanning sessions as part of the study: one before beginning the IIPT program, and a second within one week of completing the 4-6-week IIPT program. Prior to each MRI session, the sensory filament task was calibrated by starting with a very soft filament applied to the palmar surface of the left thumb and asking participants to report pain on an 11-point numeric rating scale (NRS). The research assistant moved to subsequently stronger filaments, and the filament chosen for use during fMRI was the first filament rated greater than 5/10 pain intensity. Participants were then positioned in the scanner and the anatomical and functional MRI images were collected in this order: localization scan, MPRAGE (Magnetization-Prepared Rapid Acquisition Gradient Echo), BOLD (Blood Oxygen Level Dependent) Picture Task (Run 1), BOLD Sensory Filament Task (Run 1), BOLD Picture Task (Run 2), BOLD Sensory Filament Task (Run 2). The Picture fMRI task involved passive viewing of emotional and pain-related images including some that have been widely used in previous neuroimaging research (e.g., the International Affective Picture Set, Human Facial Expressions [19]). The Sensory Filament fMRI task involved systematic external application of somatosensory stimulation (pressure) with the filament to the participant’s palmar area near the thumb in alternating blocks. The entire imaging procedure took approximately an hour. MRI technicians and study staff administered the scanning protocol. Trained research staff were responsible for the systematic external application of sensory input.

### Imaging acquisition parameters

Imaging data were collected on a 3-Tesla Siemens PRISMA scanner with a 20-channel head and neck receive coil. Participants were positioned head-first, supine in the scanner. A 3D T1-weighted MPRAGE sequence was obtained with the following parameters: Number of sagittal slices = 192; Slice thickness = 0.9 mm; Gap = 0 mm; In-plane resolution = 0.9 × 0.9 mm; Field of view = 256 × 256 voxels; TR = 2.0 seconds; TE = 2.32 ms; Flip angle = 8 degrees. Four BOLD echo planar imaging (EPI 2D PACE) sequences were obtained with the following parameters: Number of axial slices = 41; Slice thickness = 3.00 mm; Gap = 0.75 mm; In-plane resolution = 2.30 × 2.30 mm; Field of view = 94 × 94 voxels; Number of volumes = 116; TR = 3.27 seconds; TE = 30 msec; Flip angle = 90 degrees.

### fMRI task

Two tasks were conducted during fMRI scanning: a Sensory Filament Task and a Picture Task. During the Picture Task, participants viewed a slide show of still pictures while in the MRI. The pictures related to physical pain, emotional pain, and neutral pictures of office supplies as the control. The Picture Task sequences lasted for 6 minutes and 20 seconds in a block design. In each block, 10 pictures from each condition were displayed sequentially. Each picture was displayed for 2.5 seconds with a 0.5 second gap between pictures. Each BOLD sequence began with a black screen for 30 seconds. Blocks for the conditions alternated and were repeated three times for each sequence with control blocks between each condition (i.e., Control, Physical Pain, Control, Emotional Pain, Control, Physical Pain, Control, Emotional Pain, Control, Physical Pain, Control). During the Sensory Filament task, the filament was applied to the participant’s palm near the thumb by the researcher in blocks of 30 seconds alternating between Rest (Control) and Pain for 6 minutes and 20 seconds. The researcher was prompted with a visual cue to apply the filament for the variable duration of each Pain block. The timing of the Pain stimulus onset and offset is included in Supplementary Table 1.

### Data Processing

Imaging data were processed in the Analysis of Functional Neuro Images (AFNI) software package [20, 21]. Unprocessed DICOM images were converted to NIFTI format using dcm2niix [22]. Images were visually inspected for motion, artifacts, or other incidental findings. MPRAGE images were normalized to standard Montreal Neurologic Imaging (MNI152) space using the SSWarper program in AFNI. Preprocessing for functional images was conducted using afni_proc.py. Steps included de-spiking, outlier detection, motion correction, alignment to the SSwarper structural image, normalization to MNI space, and smoothing using a 4 mm gaussian kernel. Volumes with motion greater than 0.3 mm between frames or determined to be outliers were censored and removed from further analysis. Preprocessed images and the output of AFNI’s QC program were inspected for accuracy and quality using a combination of visual inspection and assessment of quantitative metrics [23, 24]. No datasets were removed for data quality issues.

### Statistical analysis

Change in pain and other questionnaire measures was assessed in SPSS 25 (IBM, Armonk, NY) comparing scores before and after the IIPT program with repeated measures analysis of variance (ANOVA). For ANOVA, significance level was set at *p* < .05.

Regression analysis was performed in AFNI in a two-stage model. A regression model was run on each participant’s data separately for the Sensory Filament Task and the Pain Picture Task as part of the preprocessing script, and then beta weights for each condition were included in group analyses using AFNI’s 3dttest++ program to compute two-sample t-tests and correlation analyses with behavioral measures.

The regression analysis for the Sensory Filament Task included conditions for Pain with Rest blocks as the Control. The regression analysis for the Picture Task included conditions for Physical Pain Pictures and Emotional Pain Pictures and office supplies were used as the Control. Contrasts of Physical Pain Pictures – Control; Emotional Pain Pictures – Control, Physical – Emotional Pain Pictures, and the main effect of Physical and Emotional Pain Pictures were calculated.

Change in brain activation during the Physical Pain Experience (Sensory Filament), Physical Pain Pictures, and Emotional Pain Pictures after completing the IIPT program was correlated with change in the questionnaire measures of Catastrophizing and Disability (FDI) using 3dttest++ in AFNI. For this exploratory pilot study, brain activations reaching a voxel-wise *p* <.01 and a minimum cluster size of 25 voxels are reported.

## Results

### Participants

18 youth (1 male, 17 female) were enrolled in the study, with ages ranging from 14-18 years (M_age_ = 16.20 years, SD = 1.31). Most participants identified as White (72.20%; n=13). The remaining participants identified as Black or Biracial (22.20%; n=4) and Hispanic Non-White (5.60%; n=1). Demographics are summarized in Table 1.

**Table 1.** Demographics.

| **Table 1. Demographics** | *M* | *SD* |
| --- | --- | --- |
| Age (years) | 16.20 |  |
| Gender (percent) |  |  |
| Male | 5.60 |  |
| Female | 94.40 |  |
| Ethnicity (percent) |  |  |
| White | 72.20 |  |
| Black or Biracial | 22.20 |  |
| Hispanic Non-White | 5.6 |  |
| Pain Type (percent) |  |  |
| Widespread | 88.90 |  |
| Central | 11.10 |  |
| Pain Duration (Weeks) | 43.60 | 27.28 |
| IIPT Duration (Weeks) | 4.30 | 1.41 |

*Note:* The age of participants ranged from 14-18 years with an average of 16.20 years.

### Changes on self-report measures

Results of the repeated-measures ANOVAs for change in Average Pain (VAS), Catastrophizing, FDI, FOPQ-C, and FOPQ-P scores revealed that each decreased after the IIPT program. Average Pain ratings also decreased, but this change did not reach statistical significance. Statistics are reported in Table 2. Individual participant’s change on Catastrophizing and Disability are presented in Figure 1A.

**Figure 1.**
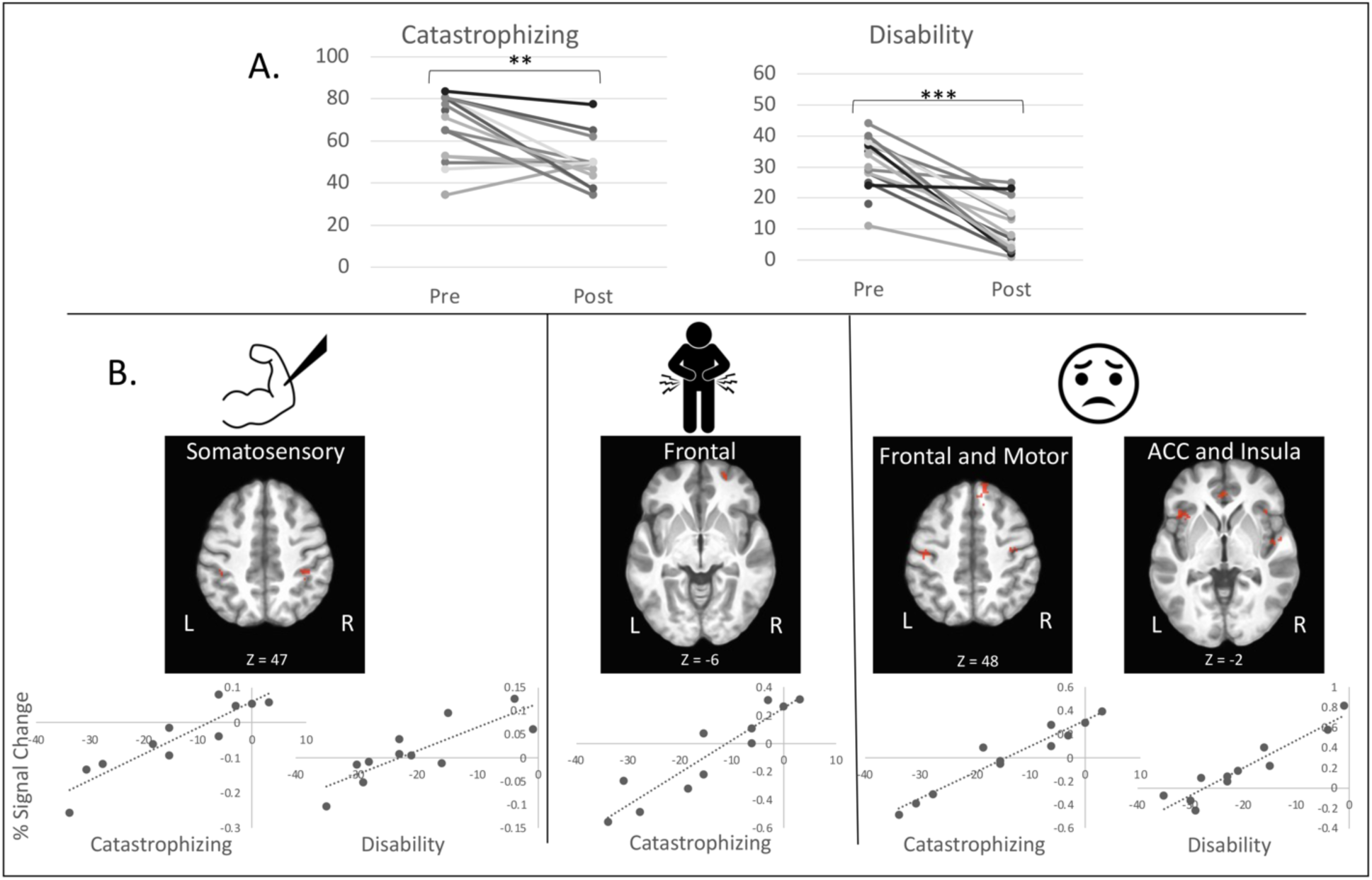
A. Line graphs of each participant’s Pre and Post IIPT program scores for the Pain Related Cognitions Questionnaire for Children (PRCQ-C) Catastrophizing subscale and the Functional Disability Inventory (FDI). Nearly all participants’ Catastrophizing and Disability improved after the program. ** = p<.01; *** = p<.001. B. Correlations between change in Catastrophizing and Disability scores and change in brain activation during each task: sensory filament pain experience (left panel), viewing physical pain pictures (middle panel), and viewing emotional pain pictures (right panel). Clusters exceeding the statistical threshold for significance of voxel-wise p<.01 and a cluster size more than 25 contiguous voxels are indicated in red indicating positive correlation. The experience of physical pain, viewing physical pain, and viewing emotional pain were all associated with improvements in Catastrophizing and Disability, and were positively correlated with reductions in activation in different brain networks: experience of pain in sensorimotor cortex; viewing physical pain in frontal cortex; and viewing emotional pain in frontal cortex, motor cortex, anterior cingulate, and anterior insula.

**Table 2.** Pediatric Rehabilitation for Amplified Pain Syndrome (RAPS) Questionnaires.

|  |  |  |  |  |  |  |
| --- | --- | --- | --- | --- | --- | --- |
|  | *M (SD) Pre* | *M (SD) Post* | *F* | *df* | *p* | *η<sup>2</sup>* |
| Average Pain (VAS) | 63.53 | 55.53 | 3.28 | 1,14 | .09 | .19 |
| Catastrophizing | 65.70 | 48.88 | 14.27 | 1,14 | .002** | .51 |
| FDI | 32.27 | 11.00 | 61.64 | 1,14 | <0.001*** | .82 |
| FOPQ-C | 51.25 | 29.50 | 17.08 | 1,11 | .002** | .61 |
| FOPQ-P | 36.42 | 23.33 | 8.04 | 1,11 | .02* | .42 |

*Note.* \* < .05. \*\* < .01. \*\*\* < .001.
*Abbreviations:* VAS = Visual Analog Scale
*FDI* = Functional Disability Inventory
*FOPQ-C* = Fear of Pain Questionnaire – Child.
*FOPQ-P* = Fear of Pain Questionnaire – Parent.

### Imaging Results

The repeated-measures ANOVA for the Sensory Filament task showed significant reduction in brain activation in the right thalamus and right caudate nucleus to the physical experience of pain with the IIPT program. Additionally, several brain regions, including right dorsolateral prefrontal cortex, right temporal cortex, left insula, and supplemental motor area/medial precentral cortex, were differentially modulated after the IIPT program in their responsivity to Physical versus Emotional Pain Pictures (Supplementary Figure 1). Overall, brain activation to Physical Pain Pictures was greater than Emotional Pain Pictures in these regions before the program, and activation to Emotional Pictures increased after the program, whereas activation to the Physical Pain Pictures decreased or did not change.

While we observed focal midbrain changes in responses to the physical experience of pain, the correlation analyses with Catastrophizing and FDI revealed several additional regions had changes in activation to the pain experience that were associated with improvements in catastrophizing and disability. Regions of the brain where program-associated modulation of brain response was associated with clinical improvement varied by task: the experience of physical pain during the sensory filament task was primarily associated with change in activation in the somatosensory cortex; the experience of viewing others in physical pain was associated with change in frontal regions; and the experience of viewing others in emotional pain modulated anterior cingulate and anterior insula – regions often associated with self-referential processing and interoception – as well as primary motor and frontal areas (Figure 1B).

Activation to painful stimulation in the right caudate, right mid-cingulate, and postcentral gyrus bilaterally was positively correlated with catastrophizing, so that greater reductions in catastrophizing were associated with greater decreases in brain activation in these regions after the IIPT program. Greater decreases in activation in right inferior parietal lobule and bilateral postcentral gyrus were positively correlated with greater reduction in disability scores on the FDI, whereas negative correlations, or greater increases in activation in right supplementary motor area, left precentral gyrus, left hippocampus, right middle occipital cortex, and left superior frontal gyrus were associated with greater reduction in disability scores.

Improvements in catastrophizing and FDI were also associated with several brain regions’ activation to Physical and Emotional Pain Pictures. Like the findings for the experience of pain, positive correlations, or greater reductions in activation to Physical Pain Pictures in right dorsolateral prefrontal cortex, right superior temporal cortex, and right inferior temporal cortex were associated with greater decreases in catastrophizing following the IIPT program (Figure 1B). Emotional Pain Pictures activations also revealed positive correlations with catastrophizing across many regions of the cortex, including primary motor cortex, temporal cortex, primary visual cortex, and superior frontal cortex. Greater reductions in catastrophizing were associated with greater decreases in activation in these brain regions following the IIPT program. Both positive and negative correlations were found between change in disability scores and activation to Emotional Pain Pictures. Positive correlations, or decreases in activation, were correlated with reductions in disability in bilateral insula, anterior cingulate, and superior temporal cortices. Negative correlations, or increases in activation, were correlated with reductions in disability in left middle frontal, right mid cingulate, right cuneus, and bilateral cerebellum. Locations and spatial extent of regions surviving thresholding for all contrasts are in Supplementary Table 2.

## Discussion

We assessed changes in brain function associated with IIPT in adolescents with widespread chronic pain using longitudinal neuroimaging. We hypothesized that improvements in clinical pain outcomes would be associated with brain activation to the experience of physical pain, and while observing another person experiencing pain, both physical and emotional. Specifically, we tested the hypothesis that there would be observable changes in the adolescent brain’s ability to distinguish between experiencing physical and emotional pain – a key component of the IIPT program.

As anticipated, we observed strong reductions in clinical pain outcomes with the IIPT program. These treatment programs are well-established for treating pediatric pain and are widely used in this population. Replicating this finding, while not novel, is critical for the interpretation of the imaging outcomes.

We observed reduced brain activation in the right thalamus and right caudate nucleus to the physical experience of pain following the IIPT program. This finding is in line with previous studies that found greater changes in the descending pain pathway following behavioral and pharmacological treatment [25–27] and may indicate a change in the top-down modulation of the stimulation from painful to nonpainful. The thalamus and caudate have been identified as key regions in the neural signature of physical pain that is sensitive to pharmacological-induced anesthesia [9]. The thalamus specifically has been implicated in chronic pain in adults and may be related to arousal during painful stimulation [28]. Somewhat surprisingly given the change in overall pain scores, we did not observe a change in other regions in this well-defined pain network with the treatment, such as the somatosensory cortex or insula which are also sensitive to physical pain severity [29]. This could be because the sensory filament only provided one type of pain stimulus (pressure) in one area of the body (palm), whereas the desensitization treatment program was broader and intended to treat widespread pain throughout the body. Our findings differ somewhat from other studies that did find differences in pain response, however those studies included children with Complex Regional Pain Syndrome (CRPS) and tested pain in the smaller area of their body affected by chronic pain, whereas the participants in our study had widespread pain [30–32].

Our examination of brain response to physical and emotional pain pictures revealed changes in several regions of the prefrontal, insular, and motor cortices with treatment. These regions are frequently observed in studies of pain processing and have been shown to have greater activation and connectivity in people with chronic pain [26, 33]. Treatment effects in these regions have also been observed; prefrontal connectivity within the default mode network and salience network have been shown to normalize with spontaneous improvement in children with CRPS [30, 32]. In our study, we hypothesized that youth with widespread chronic pain would improve their discrimination of physical pain and other types of distress with treatment, as this is a key component of the treatment exposure. Additionally, emotion discrimination, labelling, and acceptance are also a primary focus of the counseling typically included in IIPT for these youth. Therefore, we hypothesized that brain activation to pictures of physical and emotional pain would be more similar before the treatment program and diverge following the program. Our imaging findings support this hypothesis, and the greater change in response to emotional pain pictures may be related to the focus on emotions in our treatment program.

Additionally, changes in clinical outcomes related to pain – catastrophizing and disability – were correlated with changes in prefrontal, insular, sensorimotor, and limbic activation. While nearly all participants improved on both catastrophizing and disability with the treatment program, some had greater improvement than others, and this distribution was reflected in brain activation during the experience of pain and while viewing the pain pictures. Chronic pain among youth is strongly associated with protective mechanisms, including guarding emotions and fear avoidance. Increases in brain activation or connectivity in chronic pain could be interpreted as learned protective functions. Indeed, fear response is also modulated in chronic pain as has also been observed in other studies [34]. When interpreted in this way, these changes in brain activation linked to clinical outcomes may be the result of less need for protective function as the condition improves. These results provide further support for the link between pain experience and these clinical outcomes, as well as highlighting the sensitivity of neuroimaging as a tool for understanding potential mechanisms that may underlie IIPT treatment programs, or for tailoring individual treatment plans [28, 35], although self-report and subjective measures remain critical [36].

### Limitations

While the sample size in the current study is relatively modest, the novel and unique features of the project are compelling. Due to the pilot nature and statistical threshold used in this study, brain activation results should be interpreted cautiously and require further replications in the future.

The sample in the current study was almost entirely female. While we know that chronic pain conditions are more common in female youth than males, the potential effects of estrogen/testosterone on pain are not yet well understood. Future studies should include greater gender diversity among participants. Finally, we do not have long-term follow-up to know how lasting these brain changes may be. Future studies should follow participants for months to years after the intervention to determine long-term effects of treatment.

## Conclusion

In summary, youth with widespread chronic pain experienced expected improvements in functioning, fear of pain, and catastrophizing. Results show how changes in the experience of pain and the brain responsiveness to physical and emotional pain images correlate to pain and function and suggest potential brain-based mechanisms that may underlie treatment efficacy.

## Data Availability

All data produced in the present study are available upon reasonable request to the authors.

## Acknowledgements

The authors thank the participants and their families for their willingness to participate in the research project. This work was supported by a CTSA grant from NCATS awarded to the University of Kansas Medical Center for Frontiers: The Heartland Institute for Clinical and Translational Research (# UL1TR000001). The contents are solely the responsibility of the authors and do not necessarily represent the official views of the NIH or NCATS. Competing interests include funding from the Cystic Fibrosis Foundation and Multiple Sclerosis Society.

---

**Supplementary Figure 1.**
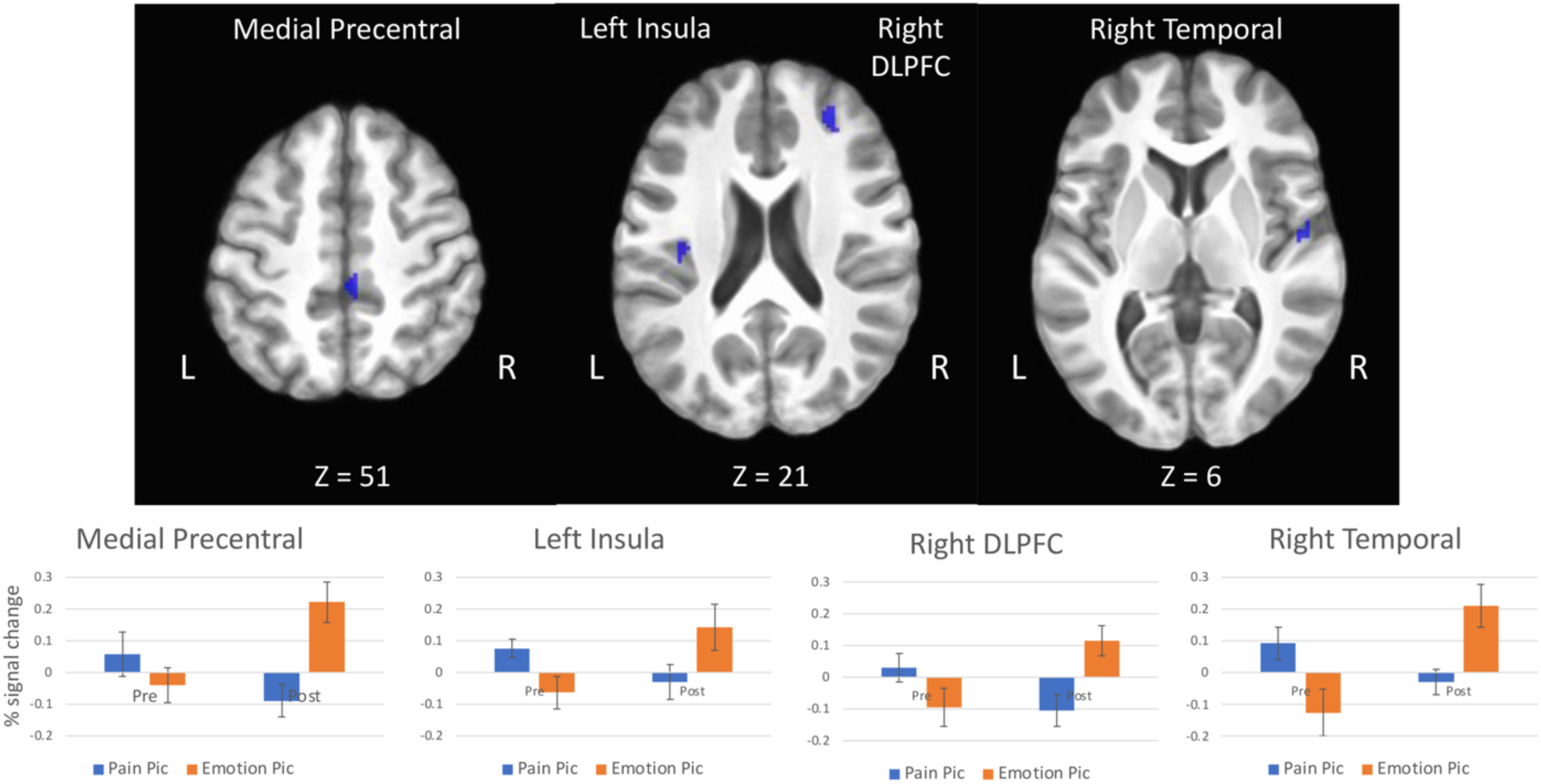
Brain activation to physical and emotional pain pictures was altered following the IIPT program. Statistical maps display results of the ANOVA for the interaction of picture type by time. Clusters exceeding the statistical threshold for significance of voxel-wise p<.01 and a cluster size more than 25 contiguous voxels are indicated in blue. Bar graphs display mean percent signal change within each region for each condition across all participants. Error bars represent standard error. Across these four brain regions, activation to physical and emotional pain pictures was more similar before the IIPT program, and activation to emotional pictures increased following the program.

