## Supplementary Table 1 for "Pediatric Neural Changes to Physical and Emotional Pain After Intensive Interdisciplinary Pain Treatment: A Pilot Study"

**Supplementary Table 1. Sensory Filament Task Time Blocks.**

|  | *On* | *Off* |
| --- | --- | --- |
| Block 1 | 4905 | 25095 |
| Block 2 | 8175 | 21825 |
| Block 3 | 3270 | 26730 |
| Block 4 | 9810 | 20190 |
| Block 5 | 6540 | 23460 |
| Block 6 | 3270 | 26730 |
| Block 7 | 8175 | 21825 |
| Block 8 | 9810 | 20190 |
| Block 9 | 6540 | 23460 |
| Block 10 | 4905 | 25095 |
| Block 11 | 8175 | 21825 |
| Block 12 | 3270 | 26730 |
| Block 13 | 9810 | 20190 |

*Sensory Filament Task Time Blocks are shown in milliseconds within each 30 second block.*
