## Supplementary Table 2 for "Pediatric Neural Changes to Physical and Emotional Pain After Intensive Interdisciplinary Pain Treatment: A Pilot Study"

**Supplementary Table 2. Imaging Results for All Tests**

*Clusters reported exceed voxel-wise p<.01 and minimum 25 contiguous voxels.*

| *Region* | *X (R-L)* | | *Y (A-P)* | | *Z (S-I)* | *#vox* |
| --- | --- | --- | --- | --- | --- | --- |
| *SENSORY FILAMENT TASK*  *Time Main Effect: Post > Pre* | | | | | | |
| Right Thalamus | 13 | | -15 | | 21 | 29 |
| Right Caudate | 11 | | 27 | | 1 | 26 |
| *PICTURE TASK*  *Image Type X Time Interaction: Physical > Emotional Pain Pictures; Post > Pre* | | | | | | |
| Medial Precentral | -3 | | 35 | | 51 | 54 |
| DLPFC | -29 | | -43 | | 21 | 51 |
| Right Temporal | -61 | | -3 | | 3 | 25 |
| Left Insula | 37 | | 15 | | 21 | 25 |
| *SENSORY FILAMENT TASK CHANGE*  *Correlation with Catastrophizing Change* | | | | | | |
| Positive Correlations |  |  | |  | | |
| Right Caudate | 7 | | 15 | | 13 | 54 |
| Left Inferior Parietal | -51 | | -43 | | 63 | 33 |
| Right Middle Cingulate | 11 | | 9 | | 35 | 27 |
| Right Postcentral | 35 | | -35 | | 47 | 27 |
| Negative Correlations |  |  | |  | | |
| None |  |  | |  | | |
| *SENSORY FILAMENT TASK CHANGE*  *Correlation with Disability Change* | | | | | | |
| Positive Correlations |  | |  | |  |  |
| Left Middle Frontal | 29 | | -1 | | 49 | 38 |
| Right Cuneus | -5 | | 81 | | 25 | 27 |
| Right Middle Cingulate | -21 | | -1 | | 43 | 30 |
| Right Parietal | -59 | | 67 | | 29 | 39 |
| Left Insula | 41 | | -21 | | 1 | 65 |
| Left Anterior Cingulate | 1 | | -41 | | -1 | 27 |
| Right Insula | -47 | | 3 | | -3 | 32 |
| Right Superior Temporal | -41 | | -11 | | -23 | 28 |
| Left Cerebellum | 39 | | 39 | | -31 | 31 |
| Left Cerebellum | -1 | | 63 | | -21 | 31 |
| Right Cerebellum | -15 | | 29 | | -25 | 34 |
| Right Entorhinal | -7 | | 7 | | -19 | 30 |
| Negative Correlations |  |  | |  | | |
| None |  |  | |  | | |
| *PHYSICAL PAIN PICTURE TASK CHANGE*  *Correlation with Catastrophizing Change* | | | | | | |
| Positive Correlations |  |  | |  | | |
| Right DLPFC | -27 | | -63 | | -11 | 48 |
| Right Superior Temporal | -63 | | -9 | | -1 | 43 |
| Right DLPFC | -29 | | -43 | | 33 | 33 |
| Right DLPFC | -41 | | -55 | | 27 | 26 |
| Right Inferior Temporal | -53 | | 31 | | -27 | 25 |
| Negative Correlations |  | |  | |  |  |
| None |  |  | |  | | |
| *PHYSICAL PAIN PICTURE TASK CHANGE*  *Correlation with Disability Change* | | | | | | |
| Positive Correlations |  | |  | |  |  |
| Left Cerebellum | -19 | | -69 | | -47 | 33 |
| Right Parahippocampal | 19 | | -21 | | -17 | 31 |
| Left Middle Occipital | -49 | | -79 | | 5 | 29 |
| Left Middle Temporal | -59 | | -41 | | -13 | 27 |
| Negative Correlations |  | |  | |  |  |
| Left Precuneus | -1 | | -67 | | 31 | 120 |
| Left Inferior Parietal | -31 | | -87 | | 47 | 64 |
| Right Supplementary Motor Area | 3 | | -21 | | 63 | 61 |
| Right Middle Cingulate | 21 | | 1 | | 41 | 47 |
| Right Precuneus | 15 | | -45 | | 33 | 45 |
| Left Inferior Parietal | -39 | | -51 | | 49 | 45 |
| Right Cerebellum | 17 | | -69 | | -39 | 42 |
| Right Superior Parietal | 21 | | -55 | | 59 | 41 |
| Left Middle Cingulate | -5 | | -41 | | 53 | 40 |
| Left Postcentral | -35 | | -5 | | 37 | 39 |
| Left Cerebellum | -17 | | -31 | | -45 | 38 |
| Right Cerebellum | 25 | | -35 | | -49 | 37 |
| Left Cerebellum | -37 | | -31 | | -33 | 31 |
| Left Postcentral | -27 | | -27 | | 75 | 30 |
| Right Hippocampus | 27 | | -37 | | -3 | 25 |
| Left Lingual | -13 | | -53 | | 3 | 25 |
| *EMOTIONAL PAIN PICTURE TASK CHANGE*  *Correlation with Catastrophizing Change* | | | | | | |
| Positive Correlations |  | |  | |  |  |
| Right Superior Frontal | -13 | | -51 | | 51 | 81 |
| Right Lingual Gyrus | -9 | | 91 | | -11 | 57 |
| Left Primary Motor | 43 | | 15 | | 49 | 48 |
| Left Inferior Parietal | 55 | | 45 | | 27 | 29 |
| Left Primary Motor | 37 | | 17 | | 41 | 29 |
| Right Supramarginal | -69 | | 51 | | 37 | 28 |
| Left Superior Temporal | 67 | | 13 | | 9 | 27 |
| Right Primary Motor | -39 | | 13 | | 45 | 26 |
| Left Primary Visual | 9 | | 63 | | 9 | 25 |
| Negative Correlations |  | |  | |  |  |
| None |  | |  | |  |  |
| *EMOTIONAL PAIN PICTURE TASK CHANGE*  *Correlation with Disability Change* | | | | | | |
| Positive Correlations |  | |  | |  |  |
| Left Anterior Insula | -41 | | 21 | | 1 | 65 |
| Right Angular Gyrus | 59 | | -67 | | 29 | 39 |
| Right Precentral | 61 | | -3 | | 47 | 39 |
| Left Postcentral | -69 | | -19 | | 17 | 37 |
| Right Insula | 47 | | -3 | | -3 | 32 |
| Right Anterior Insula | 43 | | 21 | | -9 | 31 |
| Right Parahippocampal | 7 | | -7 | | -19 | 30 |
| Right Temporal Pole | 41 | | 11 | | -23 | 28 |
| Right Middle Temporal | 49 | | -21 | | -9 | 28 |
| Left Anterior Cingulate | -1 | | 41 | | -1 | 27 |
| Right Cuneus | 5 | | -81 | | 25 | 27 |
| Left DLPFC | -55 | | 35 | | 21 | 26 |
| Negative Correlations |  | |  | |  |  |
| Left Primary Visual Cortex | -1 | | -91 | | -15 | 61 |
| Left Cerebellum | -33 | | -59 | | -39 | 39 |
| Left Precentral | -29 | | 1 | | 49 | 38 |
| Right Cerebellum | 15 | | -29 | | -25 | 34 |
| Left Inferior Temporal | -63 | | -65 | | -7 | 33 |
| Left Cerebellum | -39 | | -39 | | -31 | 31 |
| Right Cerebellum | 1 | | -63 | | -21 | 31 |
| Right Cerebellum | 9 | | -55 | | -67 | 30 |
| Right Middle Frontal | 21 | | 1 | | 43 | 30 |
| Right Hippocampus | 27 | | -35 | | 9 | 29 |
| Left Fusiform | -27 | | -9 | | -47 | 26 |
| Right Precuneus | 9 | | -59 | | 49 | 25 |
